# A genetic and clinical risk factor algorithm to aid in identifying new cases of chronic kidney disease from the general population

**DOI:** 10.1101/2024.03.21.24304689

**Authors:** Graham Rodwell, John P. A. Ioannidis, Stuart K. Kim

## Abstract

One of the biggest challenges in treating chronic kidney disease (CKD) is that 80 – 90% of people with this disease are undiagnosed, and thus do not access healthcare promptly. The problem arises because early stage CKD has no overt symptoms and the current policy is to perform diagnostic tests (e.g. glomerular filtration rate and urinary albumin to creatinine ratio) only when accompanied by risk factors such as old age, hypertension and diabetes. Genetic testing may be useful to identify those most likely to have CKD and who therefore may benefit from screening. This work describes the development of an algorithm termed RICK (for RIsk for Chronic Kidney disease) that employs a polygenic risk score for CKD plus clinical risk factors to identify people at risk. In data from the UK biobank, those in the top decile of RICK have a 4.4-fold increased risk of CKD, and about 34% of all those with CKD are included in this decile. Using RICK to selectively test those in the general population with highest risk may help in early identification of CKD and thereby facilitate early access to renal healthcare.

**Lay Summary:** One of the biggest challenges in renal health is that 80 – 90% of people with Chronic Kidney Disease (CKD) are undiagnosed, and thus do not access healthcare promptly. The problem arises because early stage CKD has no overt symptoms and the current policy is to perform diagnostic tests (e.g. glomerular filtration rate and urinary albumin to creatinine ratio) only when accompanied by risk factors such as old age, hypertension and diabetes.

This work describes the development of an algorithm termed RICK (for RIsk for Chronic Kidney disease) that employs a genetic test for CKD plus clinical risk factors to identify people at risk and who therefore may benefit from screening. Those in the top ten percentile of RICK have a 15-fold increased risk of stage 3 CKD. Diagnostic testing of the top decile would capture about 43% of the undiagnosed stage 3 CKD cases. Thus, using RICK to selectively test those with highest risk could have an immense impact on renal health by facilitating early identification of CKD and thereby enabling access to healthcare.

## Introduction

Chronic kidney disease (CKD) is defined as a depression in the filtration function of the kidneys and/or leak of serum protein into the urine.^1^ Using the widely accepted definitions of CKD advocated by the Kidney Disease Outcomes Quality Initiative and the Kidney Disease: Improving Global Outcomes CKD Work Group, the prevalence of CKD is estimated to be about 11% of the population.^2, 3^ The prevalence is increasing over time and is closely associated with age, diabetes and hypertension.^4^ Once established, CKD tends to progress regardless of cause and leads to an extensive and growing disease burden, both with associated cardiovascular illness and with the eventual outcome of end-stage renal disease, managed either with dialysis or organ transplant.

Patients with early stage CKD are typically diagnosed either by estimating their glomerular filtration rate (eGFR) or their urinary albumin to creatinine ratio (uACR), where kidney disease is defined when eGFR is below 60 mL/min/1.73 m^2^ or uACR > 30 mg/g. However, people with early stage CKD typically do not exhibit overt symptoms and screening is usually performed only on patients with risk factors, such as advanced age, hypertension or diabetes.^4^ The majority of people with early stage CKD go undiagnosed.^5, 6^ With data supporting benefit from modification of risk factors and with a growing armament of therapeutics that may slow disease progression, early identification of CKD stands to potentially have a significant impact in the management of both individual patients and healthcare populations.^7^

The heritability of CKD has been estimated from family studies to be between 30% and 75%.^8–11^ Genetic risk for CKD can be estimated using polygenic risk scores (PRS’s), which are algorithms that contain numerous DNA markers. Each DNA site contributes a miniscule amount to CKD risk, but all of the DNA sites collectively hold large amounts of information. Several PRS’s have been developed for eGFR or uACR.^12–15^

Genetic testing could benefit renal health in at least two ways. First, it could help identify new cases of CKD from those that are undiagnosed, facilitating selective screening and enabling early access to renal health care. Secondly, it could provide information that may impact management of patients already known to have CKD. Previous work has suggested that genetic testing of patients with mild CKD may aid in identifying those who are most likely to progress to kidney failure.^16, 17^

In this paper, we first examine the prevalence of cases of CKD in the UK Biobank population that have not been identified as such in electronic health records (EHR) and therefore may be undiagnosed. Then we explore how genetic screening could identify those undiagnosed or undocumented cases of CKD who may benefit from early intervention. Finally, we evaluate whether a genetic test for CKD provides information about the severity (stage) of CKD for those patients who have already been diagnosed.

## Methods

### UK Biobank Cohort

Genotype data were obtained from the v3 release of UK Biobank.^18^ The UK Biobank electronic healthcare records were available for 469,203 individuals and included data until June 2019. Genotype data were imputed centrally by UK Biobank with IMPUTE2 using the Haplotype Reference Consortium and the UK10k+ 1000GP3 reference panels ^19^. Imputed SNPs were excluded if they had an IMPUTE2 info score < 0.4. Individuals were excluded if they were outliers based on genotyping missingness rate or heterogeneity, whose sex inferred from the genotypes did not match their self-reported sex, who withdrew from participation or who were not of European ancestry. The purpose of restricting individuals to those with European ancestry is to reduce population stratification in the study. Genetic variants were excluded that failed quality control procedures in any of the genotyping batches, that showed a departure from Hardy-Weinberg of P<10^-50^ or that had a Minor Allele Frequency < .001.

### Cohorts with renal function data

The demographics and clinical characteristics of the 72,562 subjects who had both eGFR and uACR measurements are compared to those of the 243,703 who had eGFR measurements in Supplemental Table 4. The 72,562 had lower rates of female sex and higher rates of smoking than the 243,703 cohort, but the differences were relatively modest in absolute magnitude.

### CKD Definitions

Diagnosed cases of CKD (CKD-EHR) were identified based on clinical diagnoses captured in the electronic health records by UK Biobank. International Classification of Disease, Tenth Revision (ICD-10), Read v2 or Read v3 codes were used to identify cases of CKD (Supplemental Table 3). The electronic medical records originate from a diverse array of health care systems. Data from some health care systems may be incompletely represented in the UK Biobank, so some subjects may not have a diagnosis because their EHR data was not included in the UK Biobank. CKD cases based on eGFR (CKD-G) were defined as eGFR < 60 mL/min/1.73 m2. eGFR was calculated using the MDRD equation from age, sex, race and serum creatinine measurements extracted from UK Biobank.^20^ CKD cases based on either eGFR or uACR (CKD-GA) were defined as eGFR < 60 mL/min/1.73 m2 or uACR >30 mg/g. The codes used to define mild CKD and severe CKD are shown in Supplemental Table 3.

### Calculating GFR PRS scores from the UK Biobank

PRS scores for GFR and uACR and the coefficients were found by searching the PGS catalog for “GFR” or “uACR” in January 2024.^21^ The PRS scores from Yu et al., Khan et al., Sinnott-Armstrong et al. and Tanigawa et al. used cohorts that overlapped with UK Biobank 35%, 0%, 100% and 100%, respectively.^12–15^ Scores were generated for each subject using Plink2.^22^ using the coefficients from the original publication. For SNPs with missing data, an average allelic dosage was calculated based on the minor allele frequency for that SNP.

### Development of the RICK algorithm

The RICK algorithm was generated from a total of 243,703 subjects of European ancestry with complete data (seven CRFs and GFR PRS) from UK Biobank (Supplemental Table 4). The RICK algorithm was developed in R using a general linear model to fit either eGFR, CKD-G or CKD-EHR as a response with Sex, Age, BMI, blood pressure, smoking status, LDL, HbA1C and GFR PRS as predictors. The model used ten-fold cross validation, which means it was trained on 90% of the cohort and then tested on the remaining 10%. This process was repeated ten times, and then the results were averaged together. The model contains coefficients for intercept, each of the CRFs, and the PRS score (Supplemental Table 2). Values for height, weight and serum creatinine were determined by UK Biobank, and used to calculate BMI and eGFR (with the MDRD equation). Adding uACR as a risk factor to the RICK algorithm added little or no improvement to its correlation with eGFR or the AUC for CKD-EHR. Specifically, uACR was added to the RICK model and compared to RICK using 72,881 subjects (i.e. those with both eGFR and uACR data). The model with uACR had a correlation to eGFR of 0.552 and an AUC with CKD-EHR of 0.817, compared to a correlation of 0.547 and AUC of 0.796 for RICK by itself. In addition to fitting models using blood pressure and HbA1C as predictors with continuous values, alternative models were fitted using hypertension and diabetes as discrete predictors; as expected, the alternative models performed poorly compared with models using predictors with continuous values.

The performance of each PRS in predicting eGFR and CKD in the UK Biobank cohort was assessed using two independent measures: 1) correlation coefficient and variance explained and 2) area-under-the-curve. Correlation measures whether the algorithm and eGFR trend in the same direction, where 100% indicates perfect agreement and 0% indicates no relationship. Variance explained measures how well the algorithm predicts eGFR and is derived by squaring the correlation, where 100% indicates that eGFR can be precisely calculated and 0% indicates that there is no information about eGFR. Area-under-the-curve (AUC) assesses how well the algorithm classifies subjects as either CKD cases or healthy controls, where 1.00 indicates perfect reliability (sensitivity/specificity) and 0.50 indicates no more information than a random guess. CKD cases were defined either as subjects with eGFR < 60 mL/min/1.73 m2 (CKD-G) or by the electronic health records (EHR) from the National Health Service (CKD-EHR).

## Results

### Assessing the prevalence of undiagnosed CKD

The UK Biobank cohort provides an opportunity to assess the fraction of CKD cases that have no mention of a CKD diagnosis in their EHR and thus they are either undiagnosed or undocumented. There are 72,562 subjects in the cohort that had both their GFR and uACR measured as a participant in UK Biobank. GFR and uACR were used to identify people who would be diagnosed with CKD if tested within their health care system. For this paper, GFR and uACR assessed by UK Biobank were used to define four categories of CKD: 1) low eGFR (< 60 mL/min/1.73 m^2^) corresponding to stage3/4/5 CKD (CKD-G), 2) high uACR (> 30 mg/g) corresponding to proteinuria (CKD-A), 3) either low eGFR or high uACR (CKD-GA) and 4) both low eGFR and high uACR (CKD-G+A). CKD cases based on electronic health records are referred to as CKD-EHR. The percentage of cases identified by UK Biobank testing that are represented in the EHRs is 42% for CKD-G, 17% for CKD-A, 21% for CKD-GA and 70% for CKD-G+A (Table 1).

**Table 1.** Fraction of CKD cases that are diagnosed as CKD-EHR.

|  |  |
| --- | --- |
| A. Subjects with both eGFR and uACR measurements | 72,562 |
| Cases of high uACR (proteinuria; CKD-A) | 10,327 |
| Of CKD-A, number with CKD in EHR (CKD-EHR) (%) | 1,724 (16.7) |
| Cases of low eGFR or high uACR (CKD-GA) | 12,965 |
| Of CKD-GA, number with CKD in EHR (CKD-EHR) (%) | 2,727 (21.0) |
| Cases with both low eGFR and high uACR (CKD-G+A) | 1,147 |
| Of CKD-G+A, number with CKD in EHR (CKD-EHR) (%) | 802 (69.9) |
| B. Subjects with eGFR measurements | 244,671 |
| Cases of low eGFR (stages 3/4/5; CKD-G) | 9,428 |
| Of CKD-G, number with CKD in EHR (CKD-EHR) (%) | 3,961 (42.0) |

Table 2 shows the fraction of CKD-G, CKD-A, CKD-GA and CKD-G+A cases that are diagnosed as CKD-EHR for different age groups. The fractions that are diagnosed generally increase with age.

**Table 2.** Fraction of CKD-GA cases that are diagnosed as CKD-EHR as a function of age.

| AGE | <50 | 51-60 | >61 |
| --- | --- | --- | --- |
| total subjects assessed for CKD-G | 60,608 | 86,081 | 97,982 |
| subjects with CKD-G (stages 3/4/5) | 775 | 2624 | 6,826 |
| CKD-G diagnosed with CKD-EHR | 268 | 1119 | 3,846 |
| CKD-G diagnosed with CKD-EHR (%) | 34.6% | 42.6% | 56.3% |
| total subjects assessed for CKD-A/CKD-GA/CKD-G+A | 16,105 | 23,937 | 32,371 |
| CKD-A (proteinuria) | 1,838 | 3,080 | 5,409 |
| CKD-A diagnosed CKD-EHR | 132 | 383 | 1,209 |
| CKD-A diagnosed CKD-EHR (%) | 7.2% | 12.4% | 22.4% |
| subjects with CKD-GA | 1,986 | 3,713 | 7,276 |
| CKD-GA diagnosed CKD-EHR | 164 | 559 | 2,004 |
| CKD-GA diagnosed CKD-EHR (%) | 8.3% | 15.1% | 27.5% |
| CKD-G+A | 107 | 261 | 779 |
| CKD-G+A diagnosed CKD-EHR | 68 | 189 | 545 |
| CKD-G+A diagnosed CKD-EHR (%) | 63.5% | 72.4% | 70.0% |

### A genetic and clinical algorithm for CKD risk

We developed a genetic and clinical algorithm to identify those at risk for but undiagnosed with CKD. Three polygenic risk scores have been developed to predict eGFR.^12, 13, 15^ and one for uACR.^14^ Each of these PRS’s were evaluated to determine whether they should be included in the algorithm. In addition to genetics, CKD risk is affected by clinical risk factors (CRFs) such as sex, age, body mass index (BMI), systolic blood pressure (SBP), smoking status, low density lipoprotein (LDL) and glycated hemoglobin (HbA1C).^23, 24^

We first determined which of the three PRS’s for eGFR had the best performance.^12, 13, 15^ Each of the PRS’s was tested in a model including the PRS along with the seven CRF’s. Of the three GFR PRS’s that were tested, the PRS from Yu et al. performed the best (Supplemental Table 1).^15^ The model had a correlation with eGFR of 54% and explained 29% of the variance. The algorithm had AUC’s of 0.844, 0.631, 0.655, 0.818 and 0.814 for CKD-G, CKD-A, CKD-GA, CKD-G+A and CKD-EHR, respectively. The PRS from Yu et al. is hereafter referred to as GFR PRS.

Besides eGFR, uACR (>30 mg/g) is also used to diagnose CKD. We assessed the performance of the PRS for uACR in a model containing the PRS and seven CRFs. The model had a correlation with observed log(uACR) of 31.0% (95% CI; 28.6 to 33.7), explained 9.6% (95% CI; 8.2 to 11.4) of its variance and had an AUC of 0.748 (95% CI; 0.741 to 0.754) for CKD-EHR. However, the uACR PRS contributes little to the model as the seven CRFs alone also had an AUC of 0.748 (95% CI; 0.741 to 0.754) for CKD-EHR.

We then generated a model containing the PRS’s for both GFR and uACR as well as CRFs. This model had an area-under-the-curve (AUC) for predicting CKD-EHR of 0.815 (95% CI; 0.811 to 0.819). Removing the PRS for uACR had a negligible effect on the performance of the model as the AUC for CKD-EHR was 0.814 (95% CI, 0.810 to 0.818). This result indicates that the uACR PRS provided little or no contribution to predicting CKD-EHR. In summary, the final model chosen included GFR PRS and seven CRFs, but not the uACR PRS. This model is hereafter referred to as (RICK)(RIsk for Chronic Kidney disease) and its coefficients are listed in Supplemental Table 2.

The RICK algorithm is comprised of a genetic test (GFR PRS) and seven clinical risk factors. We compared the performance of the two components (genetic test vs all of the CRFs) by repeating the regression using each of the components by themselves (Table 3, Figure 1). The seven CRFs (Sex, Age, BMI, blood pressure, smoking status, LDL and HbA1C) had a correlation of 26.6%, explained 7.1% of the variance in eGFR and had AUCs of 0.720 and 0.769 for predicting CKD-G and CKD-EHR status, respectively.

**Figure 1.**
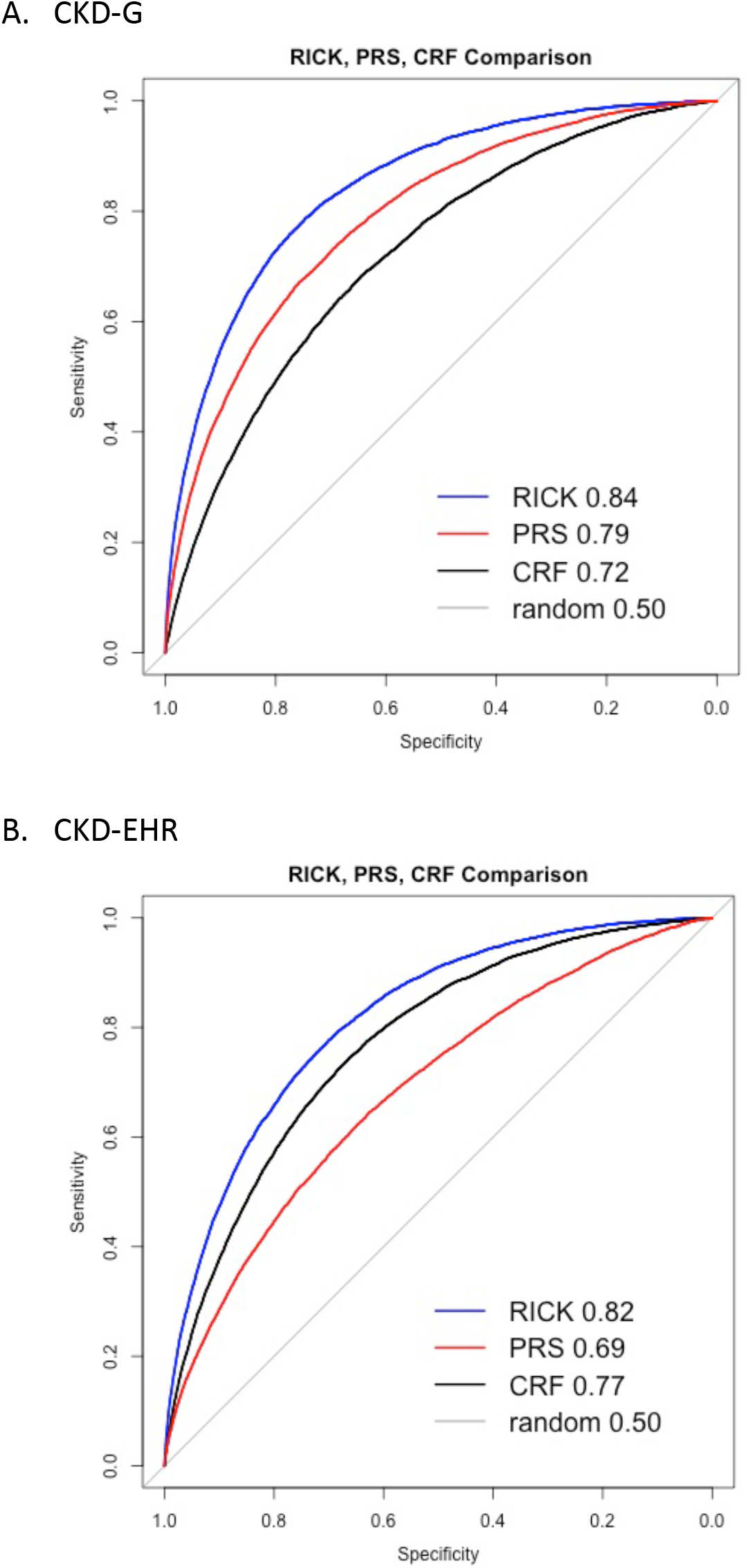
AUC plot for prediction of CKD-G (A) and CKD-EHR (B) by RICK, GFR PRS and CRFs. The AUC for each predictor is shown.

**Table 3.** Comparison of CRFs, PRS and RICK for predicting eGFR and CKD.

| Predictive Factor(s) | Performance |  |  |  |
| --- | --- | --- | --- | --- |
|  | Correlation (95% CI) | Variance explained (95% CI) | AUC CKD-G (95% CI) | AUC CKD-EHR |
| CRFs | 0.266<br>(0.141 to 0.326) | 0.071<br>(0.014 to 0.191) | 0.720<br>(0.715 to 0.725) | 0.769<br>(0.765-0.773) |
| GFR PRS | 0.465<br>(0.406 to 0.500) | 0.216<br>(0.165 to 0.250) | 0.788<br>(0.783 to 0.793) | 0.683<br>(0.678 to 0.689) |
| CRFs and GFR PRS | 0.532<br>(0.479 to 0.564) | 0.283<br>(0.227 to 0.316) | 0.843<br>(0.839 to 0.847) | 0.814<br>(0.810 to 0.818) |

CRF, clinical risk factor; GFR PRS, chronic kidney disease polygenic risk score (Yu, Jin et al. 2021); Correlation, Spearman's correlation; AUC, area under the curve.
CRFs: Sex, Age, BMI, SBP, smoke, LDL, HbA1C. All tests were performed with 244,671 subjects with complete data for CRFs and GFR PRS.

Conversely, the GFR PRS by itself had a correlation of 46.5%, explained 21.6% of the variance in eGFR and had AUCs of 0.788 and 0.683 for classifying CKD-G and CKD-EHR status, respectively. These results indicate that the GFR PRS performs substantially better than the CRFs at predicting eGFR and classifying CKD-G status, while the CRFs are better than GFR PRS in classifying CKD-EHR status. The best performance is obtained by combining both GFR PRS and the CRFs in the RICK algorithm (Figure 1).

The overall prevalence of CKD-G and CKD-EHR in the cohort used to derive RICK was 5.3% and 4.7%. RICK stratifies the population into those with either higher or lower risk for CKD. Figure 2A show the risk and prevalence for CKD-G and CKD-EHR for each decile of RICK. Subjects in the highest decile of RICK had a risk for CKD-G (18.1%) that was 15.3-fold higher than the risk among those with median RICK scores. The risk for CKD-EHR in the highest decile of RICK (16.2%) was 4.4-fold higher than the risk among those with median RICK scores. The highest decile of RICK from the UK Biobank contained 43.1% of all CKD-G cases (4,063 out of 9,428 total cases) and 34.3% of all CKD-EHR cases (3,953 out of 11,509 total). Subjects in the lowest decile of RICK had CKD-G risk of only 0.2%, 83% lower than the risk of those with median RICK scores. The lowest decile of RICK had CKD-EHR risk of only 0.07%, 81% lower than the risk of those with median RICK scores.

**Figure 2.**
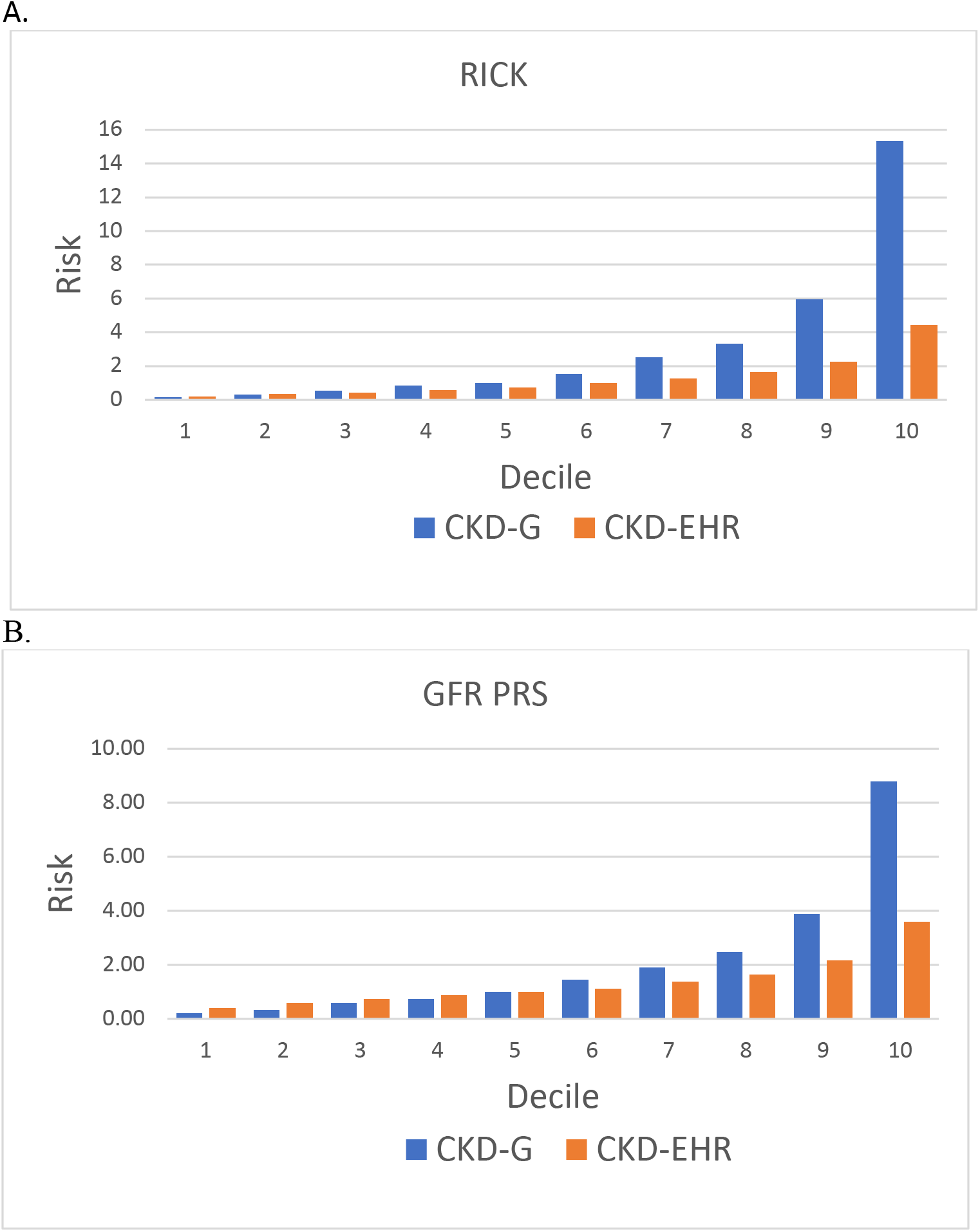
A. CKD-G or CKD-EHR risk (y-axis) for each decile of RICK (x-axis) is shown. In the highest RICK decile, the risk for CKD-G is 15.3-fold higher than the median with a prevalence of 18.1% and the risk for CKD-EHR is 4.4-fold higher with a prevalence of 16.2%. B. CKD-G or CKD-EHR risk for each decile of GFR PRS is shown. In the highest GFR PRS decile, the risk for CKD-G is 8.8-fold higher than the median with a prevalence of 15.3% and the risk for CKD-EHR is 3.6-fold higher with a prevalence of 12.6%. Risk is normalized to the median.

Figure 2B shows how GFR PRS stratifies risk for CKD. Subjects in the highest GFR PRS decile had 15.3% risk for CKD-G, 8.8-fold higher than the risk of those with median GFR PRS scores. The risk for CKD-EHR was 12.6%, 3.6-fold higher than the risk of those with median GFR PRS scores. The highest decile of GFR PRS from UK Biobank contained 3,874 cases of CKD-G, out of 9,428 total cases (41.1%); this decile had 3,076 cases of CKD-EHR, which is 26.7% of the total. Subjects in the lowest decile of GFR PRS had CKD-G risk of 0.4%, 80% lower than the risk of those with median GFR PRS scores, with a prevalence of 0.4%. The lowest decile of GFR PRS had CKD-EHR risk of 1.3%, 61% lower than the risk of those with median GFR PRS.

### Genetic testing for patients known to have CKD

We examined whether RICK or the GFR PRS alone could provide information for the CKD stage for patients already known to have CKD. In the UK Biobank cohort, there were 19,391 and 1,529 patients diagnosed with mild (stage 3 but not severe) and severe (stages 4, 5, end stage renal disease and renal transplant) CKD, respectively. If the algorithms were to provide information about the severity of CKD, one would expect that the relative number of patients with severe CKD would increase for high scores. We found that there was little change in the severe/mild ratio except at the extreme rightmost tail of the distribution. For RICK and GFR PRS, patients with scores in the highest 0.1% or 0.9% had higher ratios of severe/mild CKD (Figure 3).

**Figure 3.**
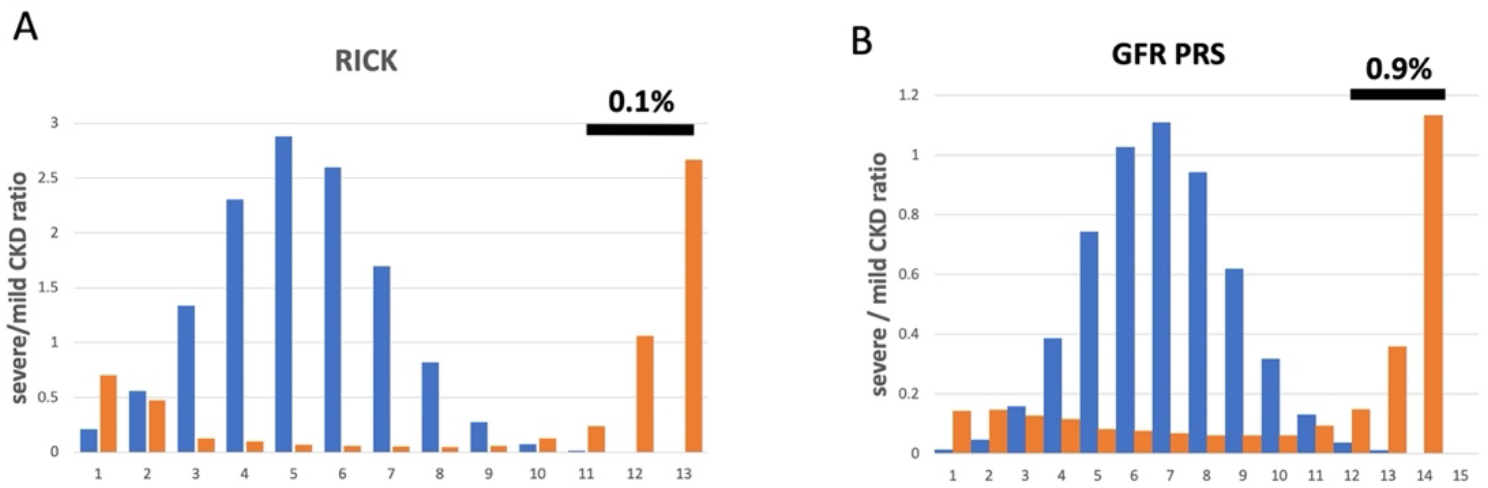
Ratios of severe/mild CKD cases (y-axis) for increasing values of RICK (A) or GFR PRS (B) are shown on the x-axis in orange. Histograms of participants used to generate the algorithms are shown in blue. The fraction of the cohort showing increased severe/mild ratios are shown.

## Discussion

This work describes the development of a genetic and clinical algorithm (RICK) that may assist in identifying people with low eGFR, one of the main criteria for diagnosing CKD. RICK could play a role in improving renal health care because CKD is underdiagnosed in the general population. Early stage CKD generally has no symptoms, and screening for CKD is typically performed only when indicated by age, hypertension, diabetes or other risk factors. As a result, a large fraction of people with CKD are undiagnosed. Previous work had estimated that 80 to 90% of CKD cases are undetected.^6, 25^ Based on the general population from UK Biobank, our data show that about 79% of the people with early stage CKD did not have CKD documented in their electronic health records (Supplemental Table 3). The high fraction of undiagnosed cases of CKD poses both a current limitation and a future opportunity to improve public renal health.

The prevalence of CKD in the general population is too low to merit eGFR measurements for everyone. The RICK algorithm could be used to identify people at risk for low eGFR. These subjects may be good candidates to have their GFR tested to find out if they have CKD, which would enable prompt first-time access to renal health care. Based on the UK Biobank cohort, the highest RICK decile contained about 43% of all CKD-G cases and 18% of subjects in this decile had CKD. A key point is that the RICK algorithm would mainly be used to identify people who should be screened for CKD.

Subsequent management would rely on well-accepted interventions to manage hypertension, glycemia, dyslipidemia and weight loss, which have been associated with reduced risk of CKD.^7, 26^ Specifically, angiotensin-converting enzyme inhibitor/angiotensin-receptor blockers,^27, 28^ inhibitors of the sodium glucose cotransporter-2,^29; 30 31^ and finerenone^32^ have been shown to slow the progression of kidney disease.^7^ Overall, preventative treatment from nephrology specialists has been shown to reduce progression of CKD compared to treatment from general practitioners.^33^

In the RICK algorithm, the GFR PRS provides substantially more information than the seven CRFs about eGFR levels and risk for CKD. Specifically, the GFR PRS explained 21.6% of the variance in eGFR while a combination of all of the CRFs explained only 8.4 percent. CRFs such as age, hypertension (blood pressure) and hyperglycemia (HbA1C) are well-accepted measures of CKD risk.^23, 24^ By analogy, GFR PRS could also be used since it is more informative.

The two main criteria for diagnosing CKD are low eGFR and high uACR. RICK is informative for low eGFR, but not high uACR. Little or no information was added by including either observed uACR or a PRS for uACR in models for predicting CKD (Methods). Thus, RICK is most useful for identifying cases of CKD due to low GFR (CKD-G) rather than high uACR.

Because it is a genetic test, GFR PRS is constant throughout life, unlike clinical risk factors that are most informative for older patients. In principle, the GFR PRS could be used to identify people with lifelong risk for CKD when they are young or middle-aged. By itself, GFR PRS has an AUC for predicting CKD-G of 0.788, which is a measurement of how well the PRS can distinguish CKD cases from controls. Although young people with a high GFR PRS would likely not yet have CKD, they would be more likely to develop CKD with age. Identification of those at risk may facilitate monitoring and early intervention.

A previous study also assessed whether a related PRS for GFR was informative in treating patients with mild CKD.^16^ In that study, patients with mild CKD were followed over a 6-year time frame during which 9.5% of patients developed kidney failure. The top decile of that PRS had a hazard ratio of 1.5 for predicting kidney failure. They also found that the PRS did not improve performance over the four-factor kidney failure risk equation,^16^ an algorithm used in research studies to predict kidney failure based on age, sex, eGFR and uACR.^34^ Our data also suggested that those with stage 5 versus stage 3 CKD did not differ in RICK and GFR PRS, except for a tiny proportion of people at the extreme tail of the distribution. Thus, RICK and the GFR PRS may be more effective at identifying undiagnosed patients that would benefit from testing rather than risk stratifying patients with established CKD.

One limitation of this study is that the cohort only included subjects of European ancestry. The GFR PRS by Yu et al. used in this study is highly similar (97% correlation) to the PRS used by Khan et al., who showed that the top 2% of their GFR PRS conveyed similar increases in CKD risk across all ancestry groups,^12, 15^ suggesting that these results may be more widely applicable. Another limitation is that RICK is not predictive for uACR, which is another criterion for diagnosing CKD. Future work may improve the PRS for uACR such that it may be incorporated into RICK. Third, about 39% of the cohort used to derive GFR PRS was from the UK Biobank, so it is possible that its performance is inflated as there is overlap between the cohorts used to derive and validate the algorithm.^15^ Fourth, the participants in UK Biobank may not accurately reflect the general population, which would affect estimates of CKD prevalence. For instance, UK Biobank participants were older than the general population. Fifth, information on electronic medical records may be incomplete. Therefore, when no CKD code is found in the EHR, this could mean either that CKD is undiagnosed or at least not documented as such. Sixth, a solid diagnosis of CKD should be defined using more than a single measurement of eGFR and/or uACR; single measurements may show fluctuation and classification of CKD based on them may lead to misclassification and both false positives and false negatives. Nevertheless, we observed high rates of undiagnosed cases regardless of the laboratory measures and their combinations used.

A major issue in using an algorithm that includes a PRS is the cost of genetic testing. Despite dramatic decreases in cost, the cost for universal population genetic screening would still be high and potentially unfavorable compared with testing for laboratory markers of CKD. This, however, will not be a barrier if the genetic information has already been obtained for other, diverse purposes, as is increasingly the case. Many millions of people have already had their genotypes determined, either through direct-to-consumer DNA testing companies or through their health care providers. As genotypes become more available, it will be possible to calculate PRS scores for many types of chronic diseases, including CKD, and thus spread the cost of genetic testing across many genetic diseases. It may thus be reasonable to use existing genetic data for the general population to identify those at risk for CKD and thereby enable selective screening and early intervention. Genetic screening would facilitate early identification of individuals at risk of CKD throughout their lifetime, long before other relevant clinical risk factor may be present. Careful decision- and cost-effectiveness analysis is needed to assess the relative utility of different screening strategies in different settings and with different cost assumptions.

A second major issue for the utility of such genetic screening is whether it can lead to better outcomes. This will need to be demonstrated in prospective studies. Nevertheless, given the potentially substantial benefit of early intervention in terms of slowing disease progression along with the growing armament of therapeutics available to impact both CKD and associated cardiovascular disease, identification of populations at risk may have a favorable impact both for individual patients as well as for policy makers managing at-risk populations.

## Supporting information

Stable1

STable2

STable3

STable4

## Disclosure Statement

SKK is CEO of AxGen, Inc.

## Data Sharing Statement

Data used in the published polygenic risk scores described in this paper are available at https://www.pgscatalog.org/ using the PGS codes shown in Supplemental Table 1. Coefficients for the RICK algorithm are shown in Supplemental Table 2.

## List of Supplementary Materials

Supplemental Table 1. Comparison of different PRS’s for eGFR and uACR.

Supplemental Table 2. Coefficients for the RICK algorithm.

Supplemental Table 3. EHR codes used to identify cases of CKD-EHR from the NHS.

Supplemental Table 4. Demographics of the cohort used to generate RICK.

## Notes

### Funding Statement

No funding was used.

### Author Declarations

The study used ONLY openly available human data that were originally located at UK Biobank (https://www.ukbiobank.ac.uk/).

