## Supplementary material for "A genetic and clinical risk factor algorithm to aid in identifying new cases of chronic kidney disease from the general population": Stable1

Supplemental Table 1. Comparison of different PRS's for predicting CKD risk.

| | eGFR | | AUC | | | | |
|---|---|---|---|---|---|---|---|
| PRS + CRFs | R | R2 | CKD-G | CKD-A | CKD-GA | CKD-G+A | CKD-EHR |
| PGS000884 | 0.54 | 0.29 | 0.844 (0.839 to 0.847) | 0.631 (0.625 to 0.638) | 0.655 (0.650 to 0.660) | 0.818 (0.805 to 0.830) | 0.814 (0.810 to 0.818) |
| PGS002237 | 0.51 | 0.26 | 0.829 (0.824 to 0.832) | 0.631 (0.625 to 0.637) | 0.652 (0.647 to 0.658) | 0.797 (0.784 to 0.810) | 0.810 (0.806 to 0.813) |
| PGS000682 | 0.47 | 0.22 | 0.807 (0.801 to 0.810) | 0.632 (0.626 to 0.638) | 0.650 (0.645 to 0.655) | 0.772 (0.758 to 0.786) | 0.800 (0.796 to 0.804) |

| | uACR | | | | | | |
|---|---|---|---|---|---|---|---|
| | R | R2 | | | | | |
| PGS001107 | 0.31 | 0.10 | ND | 0.660 (0.655 to 0.666) | 0.652 (0.647 to 0.658) | 0.721 (0.706 to 0.735) | 0.749 (0.742 to 0.755) |

eGFR, estimated glomerular filtration rate; AUC, area under the curve, uACR, urinary albumin to creatine ratio. PRS codes are from the PGS catalog (https://www.pgscatalog.org/). R and R2 are the correlation and variance explained for eGFR. CKD-G (eGFR < 60 mL/min/1.73 m2), CKD-A (uACR > 30 mg/g), CKD-GA (CKD-G or CKD-A), CKD-G+A (CKD-G and CKD-A), CKD-EHR (CKD coded in electronic health records)
