## Supplementary material for "A genetic and clinical risk factor algorithm to aid in identifying new cases of chronic kidney disease from the general population": STable2

Supplemental Table 2. Coefficients for the RICK algorithm.

| | Estimate | SD |
|---|---|---|
| (Intercept) | 159.983 | 0.353 |
| Sex | 2.105 | 0.057 |
| Age | -0.495 | 0.004 |
| BMI | -0.259 | 0.006 |
| SBP | 0.037 | 0.001 |
| smoke | 4.096 | 0.105 |
| LDL | 0.245 | 0.033 |
| HbA1C | 0.117 | 0.005 |
| GFR PRS | -29.018 | 0.108 |

Sex (F=0, M=1); BMI, body mass index (kg/m2); SBP, systolic blood pressure (mmHg); HbA1C (mmol/mol); LDL, low density lipoprotein (mmol/L); GFR PRS, GFR polygenic risk score (Yu, Jin et al. 2021). All coefficients were significant at $p < 1E-16$.
