## Supplementary material for "A genetic and clinical risk factor algorithm to aid in identifying new cases of chronic kidney disease from the general population": STable3

Supplemental Table 3. EHR codes used to identify cases of CKD-EHR from the NHS.

| ICD-10 | Description | Subjects | CKD severity |
|---|---|---|---|
| N18.0 | End-stage renal disease | 461 | Severe |
| N18.3 | Chronic kidney disease, stage 3 | 15,125 | Mild |
| N18.4 | Chronic kidney disease, stage 4 | 2,024 | Severe |
| N18.5 | Chronic kidney disease, stage 5 | 1,562 | Severe |
| M01 | Transplantation of kidney | 17 | Severe |
| Read2 | | | |
| 1Z12 | Chronic kidney disease stage 3 | 2,626 | Mild |
| 1Z14 | Chronic kidney disease stage 5 | 44 | Severe |
| 1Z15 | Chronic kidney disease stage 3A | 232 | Mild |
| 1Z16 | Chronic kidney disease stage 3B | 54 | Mild |
| 1Z1B | CKD stage 3 with proteinuria | 23 | Mild |
| 1Z1C | CKD stage 3 without proteinuria | 101 | Mild |
| 1Z1D | CKD stage 3A with proteinuria | 22 | Mild |
| 1Z1E | CKD stage 3A without proteinuria | 243 | Mild |
| 1Z1G | CKD stage 3B without proteinuria | 46 | Mild |
| 1Z1T | CKD G3aA1 - chronic kidney disease with glomerular filtration rate category G3a and albuminuria category A1 | 11 | Mild |
| K053 | Chronic kidney disease stage 3 | 242 | Mild |
| Read3 | | | |
| XaLHI | Chronic kidney disease stage 3 | 6,034 | Mild |
| XaLHK | Chronic kidney disease stage 5 | 75 | Severe |

| | | |
|---|---|---|
| | CKD-EHR total | 23,158 |

Subjects indicates the number of unique individuals with the diagnostic code from UK Biobank.
