## Supplementary material for "A genetic and clinical risk factor algorithm to aid in identifying new cases of chronic kidney disease from the general population": STable4

Supplemental Table 4. Demographics of the cohort used to generate RICK.

| | complete cases eGFR and uACR | | | complete cases eGFR | |
|---|---|---|---|---|---|
| | Female | Male | p-value[a] | Female | Male |
| Sex | 36,002 (49.6%) | 36,560 (50.4) | < 2.2e-16 | 134,787 (55%) | 108,916 (45%) |
| | yes | no | | | |
| smoke | 6,717 (9.2%) | 65,845 (90.8%) | < 2.2e-16 | 18,932 (7.8%) | 224,771 (92.2%) |
| | Average | SD | | | |
| Age | 57.64 | 8.00 | 0.97 | 56.83 | 8.01 |
| BMI (kg/m^2) | 28.29 | 5.14 | 0.94 | 27.29 | 4.66 |
| SBP (mmHg) | 138.90 | 22.06 | 0.93 | 134.66 | 21.19 |
| HbA1C (mmol/mol) | 37.10 | 8.14 | 0.96 | 35.90 | 6.36 |
| median uACR (mg/dL) | 9.35 | 119.56 | 1.00 | NA | NA |
| LDL (mmol/L) | 3.54 | 0.90 | 1.00 | 3.57 | 0.87 |
| CKD_score | 1.70 | 0.26 | 0.99 | 1.71 | 0.26 |
| eGFR | 86.59 | 16.38 | 1.00 | 86.49 | 16.33 |
| total subjects | 72,562 | | | 243,703 | |

a. Chi-squared or t-tests were used to evaluate differences between the cohorts with 72,562 and 243,703 subjects.
